# The Vitality Project: A Randomized Control Trial Comparing Effects of Qigong and Exercise/Nutrition Training on Fatigue, Emotional Health, and Stress in Fatigued Female Cancer Survivors

**DOI:** 10.1101/2022.08.18.22278965

**Authors:** Chloe S. Zimmerman, Simona Temereanca, Dylan Daniels, Cooper Penner, Tariq Cannonier, Stephanie R. Jones, Catherine Kerr

## Abstract

Cancer-related fatigue (CRF) is a common and burdensome, often long term side effect of cancer and its treatment. Many non-pharmacological treatments have been investigated as possible CRF therapies, including exercise, nutrition, health/psycho-education, and mind-body therapies. However, studies directly comparing the efficacy of these treatments are lacking. To fill this gap, we conducted a parallel single blind randomized control trial with women with CRF to directly compare the effects of Qigong (a form of mind-body intervention) (n=11) to an intervention that combined strength and aerobic exercise, plant-based nutrition and health/psycho-education (n=13). This design was chosen to determine the comparative efficacy of two non-pharmacologic interventions, with different physical demand intensity, in reducing the primary outcome measure of self-reported fatigue (FACIT ‘Additional Concerns’ subscale). Both interventions showed a mean fatigue improvement of more than double the pre-established minimal clinically important difference of three (Qigong: 7.068 +/- 10.30, Exercise/Nutrition: 8.846 +/- 12.001). Mixed effects ANOVA analysis of group x time interactions revealed a significant main effect of time, such that both groups significantly improved fatigue from pre- to post treatment (F(1,22)=11.898, p=.002, generalized eta squared effect size=.116) There was no significant difference between fatigue improvement between groups (independent samples t-test: p=.70), suggesting a potential equivalence or non-inferiority of interventions, which we could not definitively establish due to our small sample size. This study provides evidence from a small sample that Qigong improves fatigue similarly to standard exercise-nutrition. Qigong additionally significantly improved secondary measures of mood, emotion regulation, and stress, while exercise/nutrition significantly improved secondary measures of sleep/fatigue. These findings provide preliminary evidence for divergent mechanisms of fatigue improvement across interventions, with Qigong providing a gentler and lower-intensity alternative to exercise/nutrition. This clinical trial was registered with clinicaltrials.gov.

## 1.) Introduction

Cancer-related fatigue (CRF) is a common and debilitating side effect of both cancer pathology and its treatment. As many as 45% of cancer survivors report moderate to severe fatigue years after treatment cessation^1^. Fatigue is considered more burdensome and disruptive to daily life than ongoing pain, nausea, and depression^2^. Unremitting fatigue further represents a major barrier to recovering a sense of autonomy and vitality following cancer treatment, and poses an increased risk of other poor health outcomes^3–5^. As such, CRF represents a serious burden to both the patient and the healthcare system. Appropriate treatment of CRF is more pressing than ever as the incidence of cancer rises globally alongside improved survival rates of cancer patients^6,7^.

CRF is considered a multidimensional diagnosis, defined as “a distressing, persistent, subjective sense of physical, emotional, and/or cognitive tiredness or exhaustion related to cancer and/or cancer treatment that is not proportional to recent activity and interferes with usual functioning”^3^. Due to its inherently subjective nature, self-report is still the gold-standard method for measuring fatigue^3^. Many CRF patients report poorly managed fatigue symptoms that are often met with confusion and uncertainty by healthcare professionals^8–12^. This complicates early identification and delivery of effective CRF treatments, further perpetuating the burden of CRF for the patient and health system.

While there is no gold standard treatment of CRF, five major categories of treatment have been consistently studied. These include: 1.) Pharmaceuticals, 2.) Diet/nutrition, 3.) Health/Psychoeducation Approaches, 4.) Exercise and 5.) Mind-body and meditative movement approaches^3,13–17^. Pharmaceuticals have shown limited efficacy in reducing fatigue, with the possible exception of stimulants such as methylphenidate, though effect sizes are considered small and inconclusive^15,17^. Behavioral interventions and lifestyle modification such as diet/nutrition, health/psychoeducation, exercise, and mind-body approaches rather than pharmaceuticals currently show the best evidence for reducing fatigue^15^.

Nutritional approaches have shown inconclusive results for reducing fatigue, though plant-based diets appear promising^18–21^. However, long-term dietary changes can be difficult to accomplish and maintain when fatigued^18^. Psychoeducation has also been shown to be effective, either on its own or in combination with other exercise courses^15^. A recent meta-analysis indicates that exercise has the largest evidence base for fatigue reduction during and after cancer treatment^17^. However, there is considerable variation in the type of exercise utilized across these studies (eg, stretching vs strength training vs aerobic exercise). Additionally, standard exercise can also be prohibitively intense for a physically deconditioned cancer survivor struggling with fatigue, which limits adherence to exercise protocols in the short-term despite the long-term benefits ^22–24^. Together, nutrition, psychoeducation, and exercise form the core of “standard” medical advice for CRF treatment, when any is offered by medical providers^2,25^.

Mind-body approaches, which include Yoga, Mindfulness, Tai-Chi, Qigong, and relaxation techniques have also shown efficacy in reducing fatigue on meta-analysis and are increasingly studied for their multifaceted health effects across physical, emotional, and cognitive domains^26–31^. Importantly, many of these practices include a gentle, low intensity movement component which could recapitulate some of the salutary effects of standard exercise without requiring the same level of physical effort as exercise that can be difficult in a physically deconditioned cancer survivor. Non-Western mind-body movement traditions such as Tai Chi and Qigong that engage the body in a specific form of gentle movement accompanied by a meditative form of attention/awareness are commonly referred to in the research literature as “Meditative Movement” practices^13^. The specific meditative movement practice of Qigong has a rich history in Chinese medical traditions aimed at increasing a sense of vital energy^32^, and naturally incorporates movement with breath and meditative practice. Numerous different forms of Qigong and related Tai chi practice have been previously implemented in hospital settings and show promise for alleviating cancer-related fatigue and cancer-related symptoms^26,33,34^. A recent review of 13 articles examining the efficacy of Qigong for reducing cancer-related fatigue indicates that Qigong has a small-to-medium efficacy for management of fatigue, but that many studies have been limited by bias or small sample size^27,35^.

While “standard” approaches to fatigue reduction emphasize adherence to a moderate-to-vigorous exercise intervention, it is increasingly understood that lower intensity exercise has been understudied but may be uniquely suited to a fatigued population^36^. Meditative movement practices such as Tai Chi or Qigong may form an ideal version of low-intensity physical exercise that have added benefits for psychological and cognitive health due to the cultivation of meditative awareness. To this end, one recent study directly compared Tai Chi to standard moderate-to-vigorous exercise in older adults with cognitive decline and showed that both groups significantly improved cognitive function^37^. While a few studies have directly compared Qigong interventions to gentle non-meditative movements (eg line dancing, light stretching, walking)^38–41^ for fatigue reduction, to our knowledge none have directly compared Qigong to a combination of the more “standard” fatigue interventions, including lifestyle changes in nutrition and exercise. Outcomes from such comparisons are needed to principally extend standard care to include the low-metabolic Meditative Movement practice of Qigong.

To address this gap, in the current study we directly compare the Meditative Movement practice of Qigong (n=11) to a standardized combined exercise/nutrition intervention (n=13) over 10 weeks in female cancer survivors. We aimed to assess the magnitude of fatigue reduction generated by the very different physical intensity demands across the two courses. We report the results of a primary fatigue outcome endpoint as well as secondary analyses examining self-reported sleep, mood, stress, and quality of life. Though this study is underpowered to meet clinical trial significance, we provide evidence that CRF improved by similar amounts in both the Qigong and Exercise/Nutrition interventions. The Qigong group additionally reported significantly improved self-reported measures of mood, emotion regulation, and stress, while Exercise/Nutrition led to significant changes in sleep and fatigue measures. These findings may indicate both shared and divergent mechanisms of fatigue reduction in Qigong vs Exercise/Nutrition, providing a basis for further investigation in future studies.

## 2. Methods

### 2.1 Study Design

This study was a single-blind, single-center randomized control trial with a primary outcome of comparing changes in fatigue level in female cancer survivors randomized to two parallel 10-week movement-based intervention groups. All participants ranked their baseline fatigue as >3/10 in terms of its disruption of their normal daily activities or sleep quality. Participants were randomized to one of the following two intervention groups and all analyses presented include only those who completed treatment : 1.) Qigong (N=11) and 2. Pre-Train exercise/Consolidated Health Improvement Program (CHIP) nutrition + lifestyle class (N=13). Both classes met twice weekly for 2 hours and 15 minutes a class (total 4.5 hrs in-class time per week) over 10 weeks. Participants engaged in two 3.5 hour testing sessions at two different time points: prior to the classes beginning (pre), and immediately after the classes concluded (post). This study was single-blinded as participants knew their assignment in order to actively engage with the class, but study staff working on randomization, data collection, and analysis were blind to group assignment. Enrollment began in June of 2017 and concluded in August 2017. All participants completed both testing sessions and their assigned class by December, 2017. The protocol was approved by the Institutional Review Board of the local hospital where the classes were conducted. All participants gave written, informed consent prior to being randomized.

### 2.2 Participants (Table 1 for inclusion/exclusion criteria)

All participants were female cancer survivors (avg age 57.3 +/- 9.0 years) who completed cancer treatment (including surgery, radiation, and/or chemotherapy) at least 8 weeks prior to the initial phone screening. All participants met study eligibility criteria (see Table 1).

**Table 1:** Eligibility Criteria.

|  |  |
| --- | --- |
| Inclusion Criteria | Female Sex |
|  | <u>Aged 18-70 years</u> |
|  | Have completed a course of chemotherapy cancer treatment (except for ongoing treatment with Herceptin [trastuzumab] or other adjuvant therapies), AND/OR radiation treatment, AND/OR surgery, with no surgery, radiation or chemotherapy received in the past 8 weeks. |
|  | Endorse a 3/10 or above for the question “On a scale of one to ten, in the last week, how much has your fatigue interfered with your daily life, with 1 being not at all, and 10 being all the time” AND/OR “On a scale of one to ten, in the last week, how much have you felt your sleep has been disturbed or of a quality that you have not felt rested when you wake up, with 1 being not at all, and 10 being all the time” |
|  | Have a primary care or other physician |
|  | Ability to understand English |
|  | Willingness to have blood drawn |
|  | Willingness to have an EEG, EKG, and EMG taken |
|  | Ability to pass basic validated physical movement tests (e.g.: standing with feet touching for 30 seconds, twist right to left and back, hold arms out to side in air for 15 seconds, lift arms over head, moving from standing position to seated position on the floor) to verify safety for moving practice |
| Exclusion Criteria | History or current diagnosis of coronary artery or coronary heart disease |
|  | History or current diagnosis of heart attack, or heart murmur |
|  | Electrical Pacemaker Implant in heart |
|  | Peripheral neuropathy in hands |
|  | History or current diagnosis of any other MAJOR psychiatric disorder (including psychosis, or mania, or suicidal or homicidal ideation) besides depression or anxiety |
|  | Active alcohol or drug abuse |
|  | Tobacco use |
|  | Pregnancy |
|  | Ingestion of caffeine or cocoa products less than two hours from data collection |
|  | Inability to participate in resistance exercises (like Pilates) |

### 2.3 Procedure

Intervention classes began in September and ended in November 2017. A variety of recruitment methods were utilized including posting flyers, placing advertisements in local radio shows and community magazines, outreach to local oncologists, oncology nurses, cancer support groups, and local organizations, and hosting stands at local events like parades and baseball games. Interested women called in, where an initial phone screen was conducted to describe the study’s goals, methods, and time requirements and to screen for eligibility. If eligibility was satisfied, the participant was scheduled for an in-person consent session, during which the participant was informed about both the Qigong class and the Exercise/Nutrition healthy living class and told that they would be randomly assigned to either class. They were asked to indicate that they were willing to be randomized to either class and didn’t have a strong expectation of which class they would be in. If the participant consented to join the study, they were then scheduled for an initial 3.5 hour testing session.

48 hours prior to a scheduled testing session, the participant was emailed the packet of questionnaires to fill out online via REDCAP before their session (∼45 minutes). If participants were not able to fill out questionnaires online, they were asked to come to their testing session 45 minutes early to do so then. Testing sessions included multiple assessments in the allotted 3.5 hours, including measures of back, leg and arm strength, a 6 minute walk test, and venipuncture. Some participants also completed an EEG, EMG, ECG, laser doppler flowmetry, working memory task, and electrodermal skin conductance measures; all measures other than the questionnaires are beyond the scope of the current study. All testing sessions took place at the hospital clinic where the classes were conducted.

### 2.4 Randomization and Blinding

We generated a 1:1 computer-based randomization scheme using the blockrand package in R. The randomization was stratified by cancer recurrence vs. no recurrence, to ensure an equal number of participants who had experienced multiple episodes of cancer and its treatment were assigned to each group. Therefore, for each stratum (cancer recurrence vs. no recurrence), subjects were randomized to one of the two treatment groups within sequential blocks with random block sizes of 2, 4 and 6 using the blockrand function. Random block sizes were used to prevent treatment allocation anticipation. A pdf of randomization cards was created based on the output from blockrand using the plotblockrand function. To ensure blinding of the study personnel to group assignment despite having limited personnel, a Data Scientist generated multiple possible randomization sequences. A Research Assistant not otherwise involved in this study then randomly selected one of the sequences, and printed the randomization cards and sealed them in envelopes. At the completion of the baseline testing session for each new subject, the next envelope was opened and the subject was assigned to the corresponding treatment. Participants were not randomized to their intervention group until the end of their first testing session to ensure that both Research Assistants and participants were blind to their intervention group at baseline testing. Participants started their assigned intervention within three weeks of completing the baseline assessment.

### 2.5 Intervention Groups

The primary objective of the study was to compare the efficacy of Qigong meditative movement to a standard healthy living program of exercise and plant-based nutritional/health education (described below) in reducing fatigue.

Both intervention groups met for a total of 4.5 hours per week (2 classes/ wk). Both intervention groups were asked to practice the movement-based component of class at least 30 minutes a day on days without classes. Both a morning and an evening class were offered to accommodate participant’s schedules for both intervention groups.

#### 2.5.1 Qigong Intervention

The Qigong class was assembled and taught by an internationally recognized Qigong Instructor with 40 years of experience in teaching Qigong in both China and the US. The focus of the class was on the Qigong/Chinese Medicine principles of “Nourishing Life” (Yang Sheng) and drew heavily from the Hun Yun system of Qigong. Practices were selected by the instructor specifically for fatigued cancer survivors because of their prior use in China in improving energy levels and physical functioning in Cancer patients. Movements were gentle, rhythmic, and repetitive and began with seated meditation, and progressed to standing movements in place. Gentle swinging of the arms and bending of the legs was the main source of exertion. A meditative focus was cultivated throughout as participants were instructed to focus on their breath and softening their muscles throughout each posture. Throughout the first six weeks of class, new Qigong movements were sequentially introduced working up to a final sequence of 15 movements. The last four weeks of class were spent solidifying and refining the full sequence. Each class was roughly allocated into a first hour of learning a new movement, a relevant topic and discussion for the day about the purpose of the movement and related Qigong principles of healing and energy cultivation. After a fifteen minute break, the second hour was spent practicing the sequence and answering any practice-related questions. Participants were given instruction sheets for each movement as it was introduced to facilitate home practice.

#### 2.5.2 Pre-Train Pilates & Complete Health Improvement (CHIP) Intervention (Pre-Train/CHIP)

This intervention group combined two pre-existing standard healthy living-focused classes incorporating both physical exercise and general health and nutrition education. Because the focus of the study was to examine how Qigong compared to standardized health advice offered by physicians to combat fatigue - mainly exercise, dietary changes, and health education - this course included components of all three. For the first hour of each class, participants were led in a program called Pre-Train, a pilates-like exercise program designed by a local Chiropractor to help improve core and overall strength. All participants were given a manual of exercises and asked to practice on their own at home 5 days a week for 30 minutes. Practices progressed throughout the 10 weeks to include both resistance and aerobic exercise. After a 15 minute break, the second hour followed the well-validated Complete Health Improvement (CHIP) program^42,43^, a standardized nutrition and healthy living educational program which has been shown to improve multiple chronic disease metrics in patients for over 25 years. This hour consisted of instructional videos, and discussions led by a licensed hospital Nutritionist. Participants received a manual and cookbook on plant-based eating and general health principles, and were encouraged to incorporate as many plant based foods into their diet as possible. There was no discussion of any aspect of breathing or meditative focus in any part of this course.

### 2.6 Questionnaires

Baseline and follow-up testing sessions were conducted at the same hospital where the interventions took place. The online REDCap system was used to collect responses to all questionnaires as well as demographic information and background information related to age, race/ethnicity, socioeconomic status, and cancer history (see Table 2). Questionnaires were emailed to participants 48 hours before their scheduled testing session, and were completed before arrival at the testing session. Trained research assistants conducted the testing sessions. Participants were asked to abstain from any caffeine consumption starting two hours before their testing session to avoid measuring the effects of caffeine on any of the biological systems.

**Table 2:**
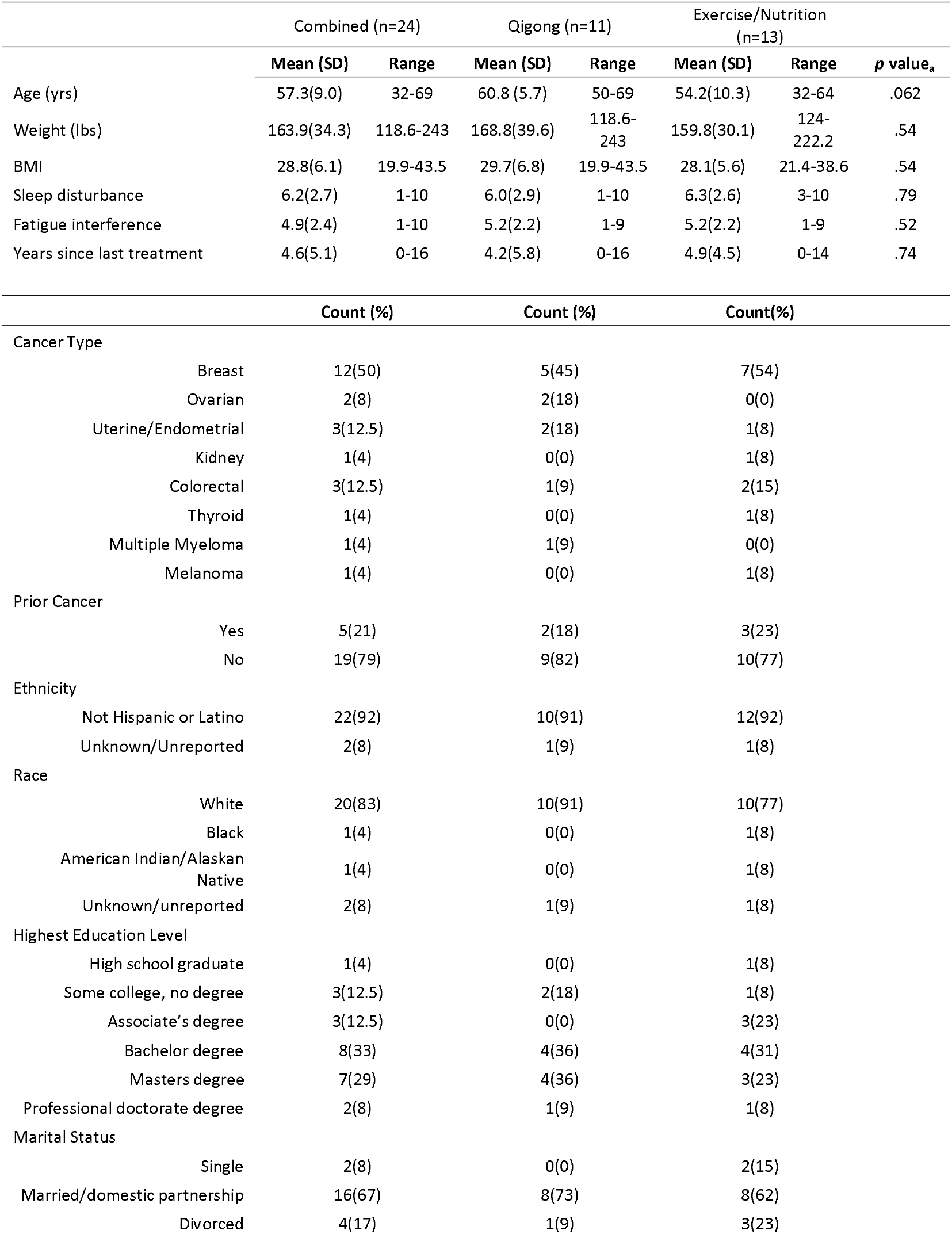
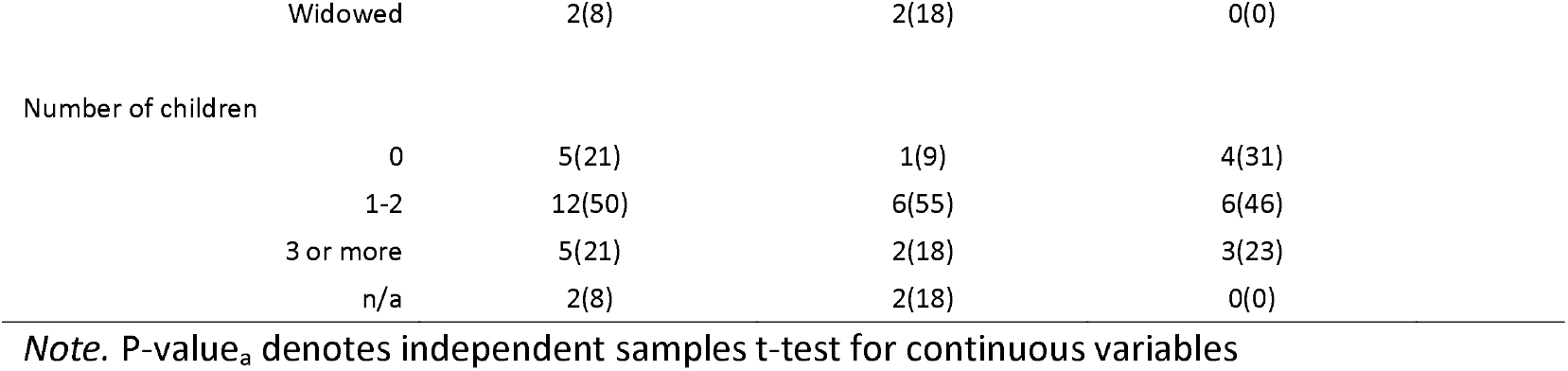
Baseline characteristics of study participants

#### 2.6.1 Primary outcome measures

The primary outcome measure was fatigue reduction. **The Functional Assessment of Cancer Therapy-Fatigue (FACIT-F)** is 41 item subscale that measures overall well-being and health-related quality-of-life (QOL) in patients with cancer and cancer-survivors to determine frequency in the past week of statements concerning physical, emotional, social, and functional well-being in addition to fatigue. The “Additional Concerns” subscale was utilized as the primary outcome measure because it directly assesses fatigue using 13/41 items on the questionnaire. For all scales, a higher score indicates improvement^44^.

#### 2.6.2 Secondary Outcome measures

Multiple questionnaires were administered to assess different aspects of fatigue experience and intervention-related improvements. These include:

##### Pittsburgh Sleep Quality Index (PSQI)

The PSQI is a 9 item questionnaire that assesses sleep habits and quality over the last month. A global score greater than 5 indicates poor sleep quality^45^.

##### Fatigue Symptom Inventory (FSI)

The Fatigue Symptom Inventory is a 14 item questionnaire that uses a likert style 1-11 scale to assess clinically meaningful fatigue, including average fatigue, fatigue interference, and fatigue frequency and severity in the last week. A higher score indicates worse fatigue^46^.

##### Multidimensional Assessment of Interoceptive Awareness (MAIA)

The MAIA is a 32 item questionnaire that assesses the frequency of certain body sensations or body related thoughts in daily life using likert style items scored 0-5. It includes subscales relating to noticing, not distracting, not worrying, attention regulation, emotional awareness, self regulation, body awareness, trusting. A higher score is better^47^.

##### Profile of Mood States (POMS)

The POMS is a 65 item questionnaire that assesses mood “right now” using different words that are scored according to subscales assessing tension, depression, anger, fatigue, confusion, vigor, and total mood disturbance on a likert style scale ranked 1-5. A higher score indicates a higher impact of that subscale^48^.

##### Difficulties in Emotion Regulation Scale (DERS)

The DERS is a 36 item questionnaire assessing general emotion regulation difficulties across 6 subscales including (1) non-acceptance of emotional responses; (2) difficulties engaging in goal directed behavior; (3)impulse control difficulties; (4) lack of emotional awareness; (5) limited access to emotion regulation strategies; (6)lack of emotional clarity. Each item is ranked likert style from 1-5. Higher scores suggest greater difficulty with emotion regulation^49^.

##### The Perceived Stress Scale (PSS)

The PSS is a 10-item questionnaire that assesses thoughts and feelings related to stress over the last month. Each item is ranked likert style from 0-4. Higher scores indicate more stress^50^.

##### Multidimensional Scale of Perceived Social Support (MSPSS)

The MSPSS is a 12-item questionnaire that assesses the amount of support someone feels from their friends, family, and significant others. Items are ranked likert style from 1-7. A higher score indicates greater social support^51^.

##### Unmitigated Communion (UC)

The UC questionnaire consists of 9 items ranked likert style from 1-5 that assess the tendency to attend to and care for others to the exclusion of the self. A higher score indicates a greater tendency to care for others over one’s self^52,53^.

##### Brief Resilience Scale (BRS)

the BRS is a 6 item questionnaire that examines the ability to bounce back after a stressor. Items are ranked likert style from 1-5. A higher score indicates greater resilience^54^.

##### Patient Health Questionnaire (PHQ)

The PHQ is a 59 item questionnaire that assesses someone’s physical and psychological symptoms over the past week using a likert style scoring of 1-3 for each item. The PHQ-9 is a subscale of 9 questions that assesses depression symptom severity. The PHQ-15 is a subscale of 15 questions that assesses severity of somatic symptoms. A higher score indicates a greater severity of symptoms^55^.

##### Short-Form-36 (SF-36)

The SF-36 is a 36-item questionnaire that assesses perceived general health and vitality in the last month as well as expectations for health. Items are ranked with radio buttons from 0-100. It consists of 8 subscales including: 1.) physical functioning, 2.) role limitations due to physical health, 3.) role limitations due to emotional health, 4.) vitality (energy/fatigue), 5.) emotional well-being, 6.) social functioning, 7.) pain, 8.) general health. Higher scores indicate better health status^56^

### 2.7 Statistical Analysis

Our primary analysis was per-protocol. A traditional intent-to-treat (ITT) analysis was beyond the point of the study if the participants didn’t complete intervention, as the goal was to assess the treatment effect of the interventions^57^. Further, the majority of people who dropped out declined to return for the second testing session making it difficult to conduct an ITT analysis. The pre-specified primary outcome measure was fatigue reduction (see 2.6.1)

#### 2.7.1 Power analysis and effect size

The power analysis was completed for the FACIT-F ‘Additional Concerns’ Fatigue subscale as the primary outcome measure. We estimated the sample size needed to achieve 80% power at the two-tailed 0.05 significance level as 66 participants completing the study (33 in each group) with an effect size of 2.5 and standard deviation of 9, using a non-inferiority margin of 3. The effect size of 2.5 and standard deviation of 9 for treatment induced changes in fatigue were estimated based on the literature^38^. The non-inferiority margin of 3 represents the minimal clinically important difference in the FACIT-F score established in previous research^58^. Power analysis was performed in R using the package Trail Size (TwoSampleMean.NIS function). Though we planned to do a non-inferiority analysis to more conclusively establish that the improvement of fatigue from Qigong practice is statistically not worse than improvement from exercise/nutrition, the low power from our small sample size per group prevented conducting this analysis in a meaningful way.

#### 2.7.2 Statistical Tests

We used mixed-effects ANOVA to assess outcome measures, along with post-hoc Holm-corrected t-tests for significant differences in ANOVA outcomes. We used Holm correction to account for multiple comparisons because it is less conservative than Bonferroni and preserves more power in our already underpowered sample^59^. Post-hoc analyses for ANOVA measures that were significant at the holm-corrected p-value below our pre-specified alpha of .05 were carried out using paired and independent samples t-tests for continuous variables. All p-values are reported as two sided tests. We report generalized eta squared effect sizes for significant ANOVA measures as this as been shown to be optimal for mixed designs^60^, computed as η_G_^2^ =SS_effect_ / (δ x SS_effect_ + ∑_measured_SS_measured_)^60,61^. We report Cohen’s D effect size for the t-test analyses of the primary outcome measure, computed as d = Mean(X_1)_ –Mean(X_2)_/pooled SD^62^. For missing data in questionnaires, we followed the protocol of the designated questionnaire for correct handling of missing data. A custom R script was used to analyze questionnaire data.

## 3. Results

### 3.1 Demographics/Baseline characteristics of participants

Demographics, baseline characteristics, and cancer history for the overall participants and broken down into groups are listed in Table 2. Participant recruitment is detailed in Figure 1, with 41 out of the initial 72 eligible participants randomized to the intervention groups (Figure 1). There were no statistically significant differences between groups for any of the continuous variables. The class retention rate was 52% for the Qigong group, and 65% for the Exercise/Nutrition group (Pre-train/CHIP), which was similar to prior studies with 8-10 week behavioral interventions^40,63^. Additionally, there were no significant group differences in any of the questionnaires at baseline (Table 3).

**Table 3.**
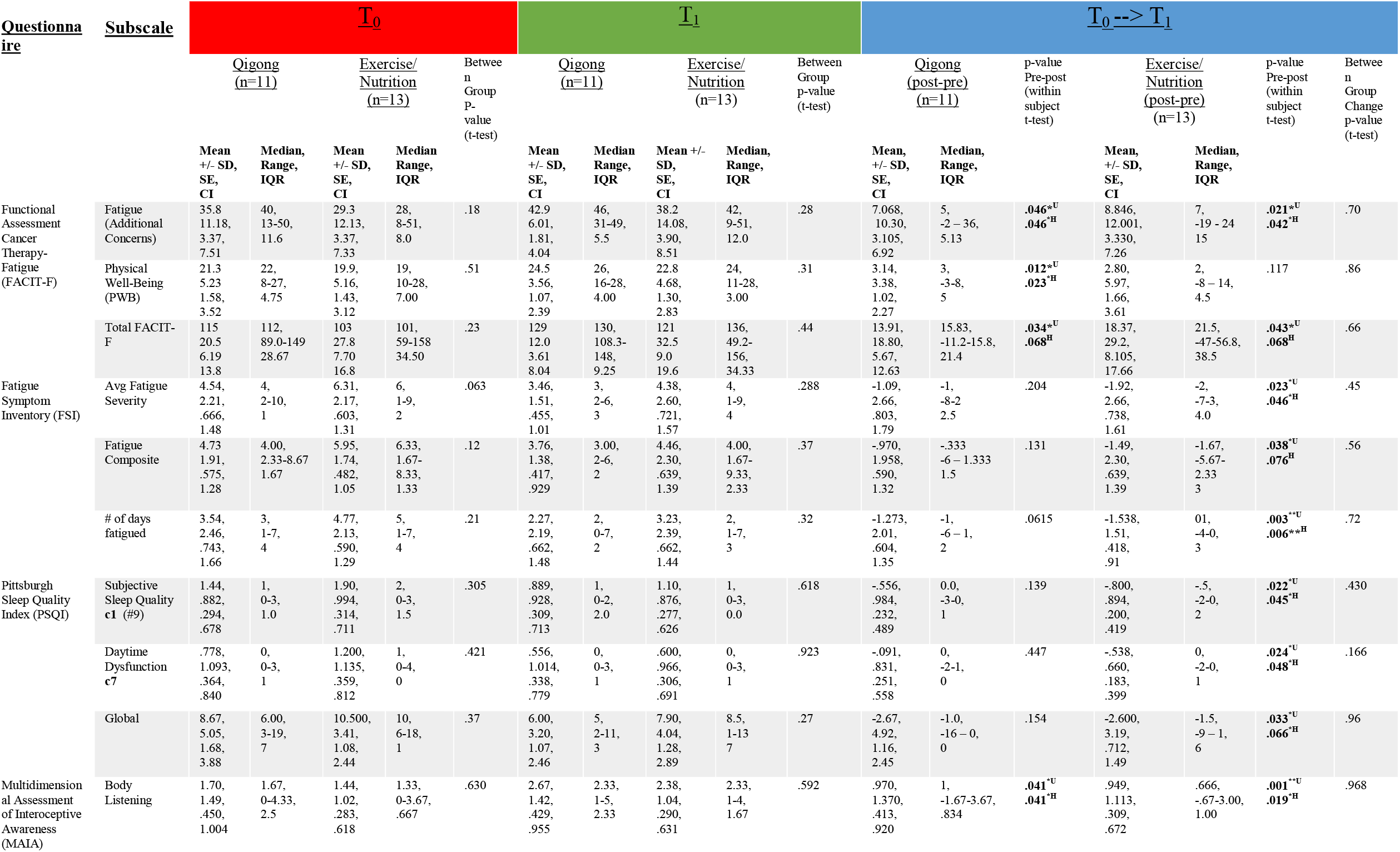
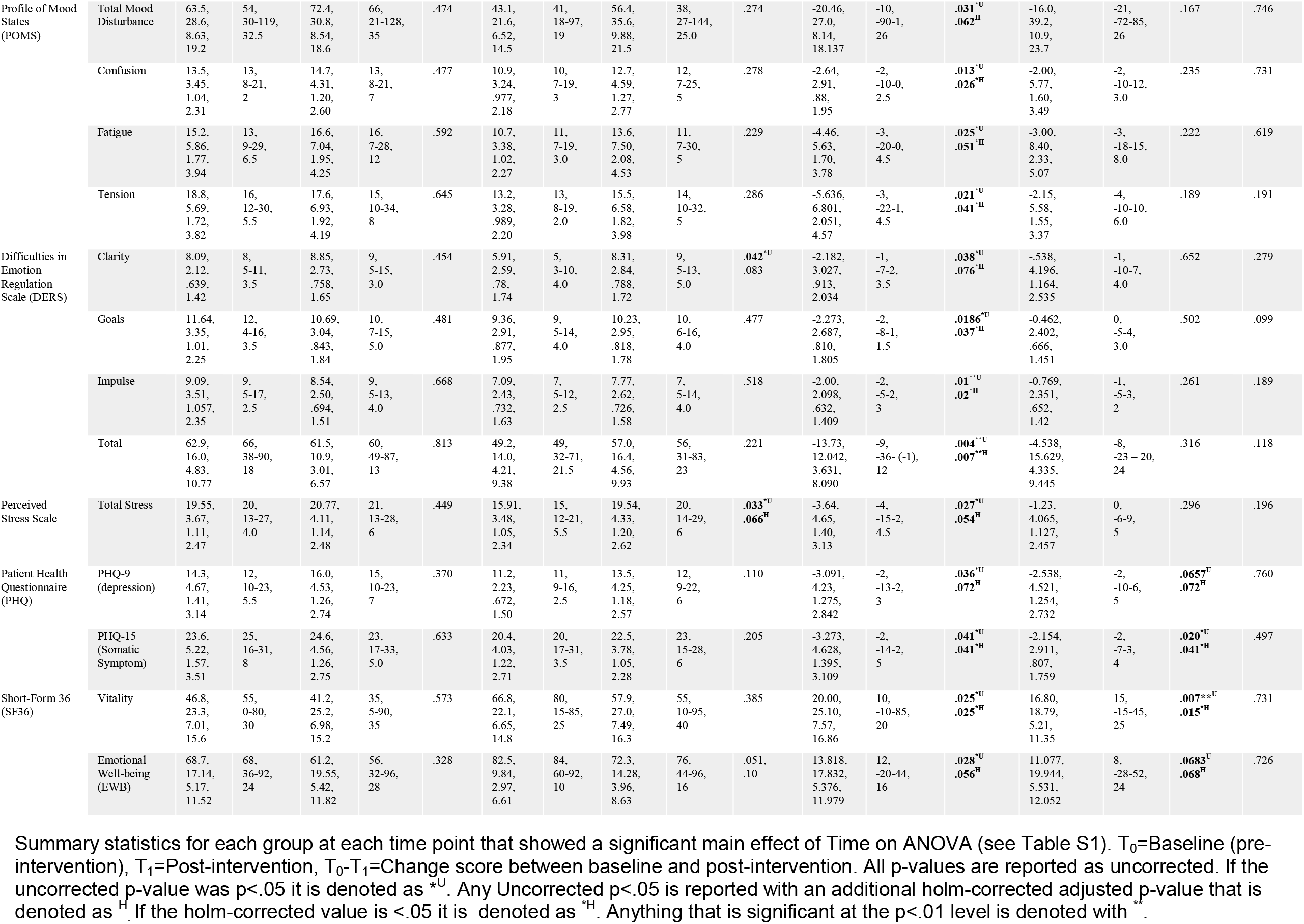
Summary of Questionnaire Subscales that Showed Significant Main Effect of Time on ANOVA.

**Figure 1.**
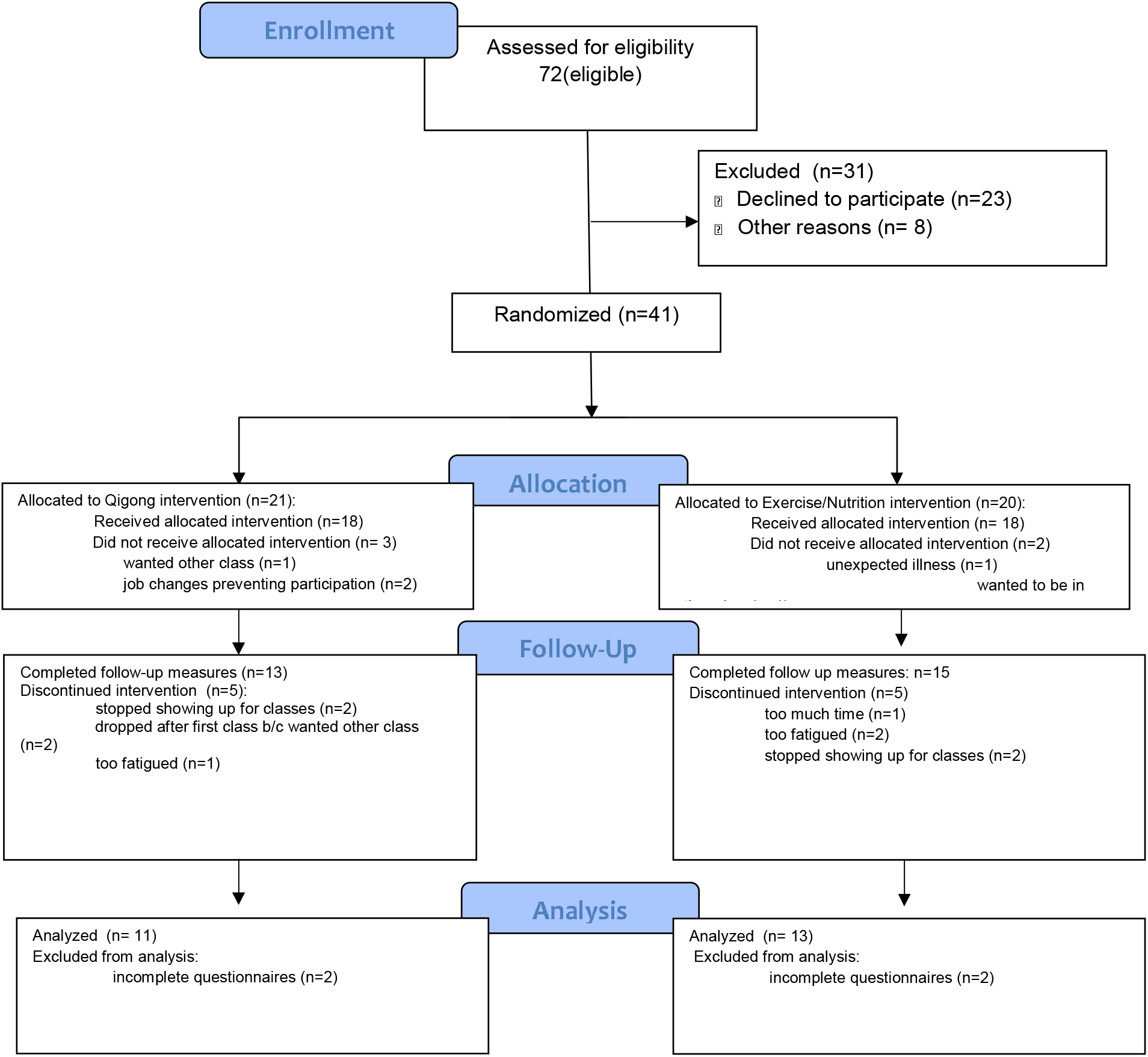
CONSORT Diagram.

### 3.2 The primary outcome fatigue measure improved in both Qigong and Pre-Train/CHIP interventions, with no between group difference

The primary endpoint of fatigue was measured with the FACIT-F “Additional Concerns” subscale and analyzed using a 2 (Group: Qigong vs Exercise/Nutrition, between-subjects) x 2 (Time: Pre vs post, within-subjects) mixed-effects ANOVA. The main effect of Time was significant, (F(1,22)=11.898, p=.002), with a generalized eta squared effect size of .116 (med-large). The main effect of Group was not significant (F(1,22)=1.915, p=.180). The interaction between Group x Time was also not significant (F(1,22)=.149, p=.704).

Exploratory post-hoc pairwise comparisons reveal that both groups improved fatigue more than the predetermined minimal clinically important difference of 3 from pre-post^58^: For the Qigong group the mean improvement in fatigue was 7.068 +/- 10.30 (t(10)= 2.28, Holm-adjusted p-value=.046, Cohen’s D=.686), while for the Exercise/Nutrition group the mean improvement was 8.846 +/- 12.001 (t(12)=2.66, Holm-adjusted p-value=.042, Cohen’s D=.738) (see Table 3). 73% of the Qigong group experienced an improvement in fatigue of greater than 3 (n=8/11) compared to 77% of the exercise/nutrition group (n=10/13). This indicates that both groups had a clinically significant effect on fatigue improvement pre-post. Further, comparison of change scores across groups does not reveal a statistically significant difference in efficacy between groups (p-value between groups=.70, see Table 3). Our findings indicate that both groups changed in similar directions and magnitude from pre-post intervention, and we could not reject the null that no difference exists between the two.

### 3.4 Secondary Outcome Measures

Given the large quantity of questionnaires assessed, many of which contained multiple independent subscales, we conducted similar secondary analyses as described above for each questionnaire. While many of the scales measuring various quality of life, fatigue, sleep, functional, emotional, social, and stress related outcomes revealed significant main effects of “Time” changes in 2×2 mixed-effects ANOVA, none of them showed any significant Group x Time interactions. Only two, the DERS ‘Clarity’ subscale and the SF-36 ‘emotional limitations’ subscale, had a significant main effect for Group in addition to Time (See Table S1). We provide a list of those scales with significant time effects along with the generalized eta squared effect size for each (Table S1).

#### 3.4.1 Post-hoc exploratory analyses

In service of further exploration of the numerous questionnaires collected, we conducted a post-hoc exploratory analysis of secondary outcome measures that had a significant “Time” effect (or “Group” effect) on ANOVA similarly to the primary outcome measure. The descriptive statistics of each scale, per group, at each time point as well as the change scores are provided in Table 3 for those scales that had a significant main effect of “Time” on ANOVA.

Though the post-hoc pairwise comparisons were exploratory given the lack of significant interaction terms on ANOVA, two patterns of note did emerge in pairwise t-tests of the change scores by group (see Table 3). First, while the significant main effect of time on ANOVA indicated that both groups improved pre-post, the t-tests of change scores across secondary fatigue and sleep outcomes indicated a significant difference mainly for the exercise-nutrition group (see **Table 3** for significant changes: FSI Avg Severity, FSI # of days fatigued, PSQI subjective sleep quality, PSQI daytime dysfunction. For trends toward significance see: FSI composite, PSQI global). Second, for the secondary measures of emotionality, mood, emotion regulation, and stress, there were significant changes pre-post primarily in the Qigong group alone (see Table 3 for Significant Changes: POMS confusion, POMS fatigue, POMS tension, DERS Goals, DERS Impulse, DERS Total. For Trends toward significant changes see: POMS total mood disturbance, DERS Clarity, DERS Nonacceptance, PSS, PHQ-9). Both groups significantly changed pre-post on multiple measures related to overall physical sensations (see Table 3 for significant changes: MAIA “Body listening”, PHQ-15, and SF36-Vitality. For trends toward significance see : FACIT-F, SF-36 emotional well-being). Though these changes appear specific to the different groups, it is important to keep in mind that there are nuances across measurement of questionnaires that assessed similar constructs. For example, while scales on the Difficulties in Emotion Regulation Questionnaire and the Profile of Mood States were significantly different pre-post for the Qigong group, the measurement of emotions via the SF--36 emotional well-being score showed a trend toward significant change in both groups (see Table 3). However, these findings may form a preliminary motivation for future studies to investigate further.

## 4.) Discussion

In this single-blind, parallel randomized control trial we demonstrated that 10 weeks of both Qigong and Exercise/Nutrition training (Pre-train/CHIP) significantly improved fatigue pre-post intervention. Both groups improved mean fatigue scores pre-post more than double the pre-established minimal clinically important difference of three^58^ (see Fig 2), with over 70% of participants in each group reporting a difference of three or more. Our results indicate that the Exercise/Nutrition group demonstrated significant improvements in self-reported secondary measures of sleep and fatigue, while the Qigong group showed significant improvements in secondary measures of emotional health and stress. The lack of statistical difference between groups on ANOVA suggests a possible equivalence of the interventions across many domains, but requires a higher-powered study to definitively assess. Establishing equivalence would have important clinical consequences, as Qigong constitutes a gentler, lower-intensity movement practice than vigorous exercise, and as such may be an effective alternative for those struggling with exercise or could be incorporated into polymodal fatigue interventions to help build stamina required for exercise adherence. Here, we provide effect sizes and descriptive statistics from our sample to help facilitate such future studies. We also include a discussion of the initial development of a manualized Qigong course for hospital based settings according to the NIH stage model.

**Figure 2.**
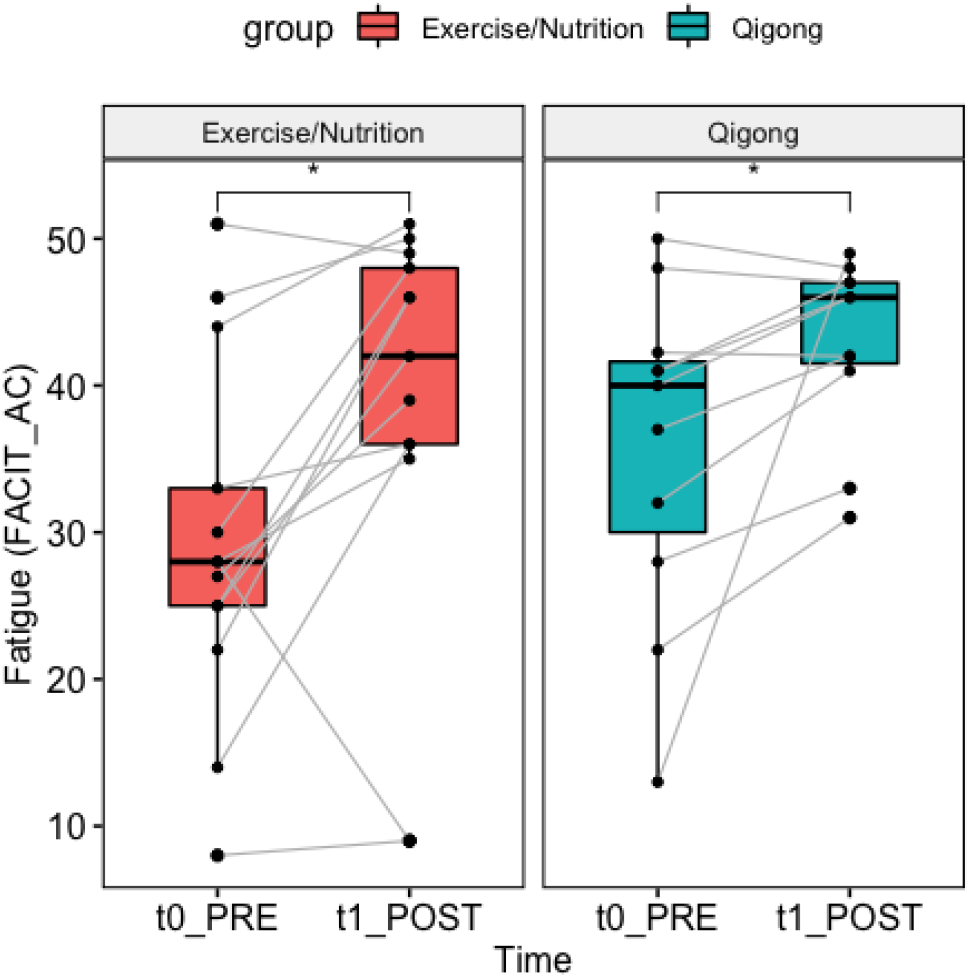
Boxplot of Fatigue Change by Group Results for fatigue change from pre-post for the primary outcome measure of fatigue, measured by the FACIT ‘Additional Concerns’ Subscale. The change from pre-post for each subject is denoted by the gray lines. * indicates a significant difference at the holm-corrected p<.05 level on paired t-tests.

### 4.1 Advantages of the study design and intervention groups compared to prior studies

A primary contribution of this study’s design was the direct comparison of Qigong to Exercise/Nutrition. While a few prior studies have compared Qigong and exercise, such studies utilized either light stretching (1 hr class, 2x/wk, 12 wks)^38^ or line dancing (mild-moderate intensity, 90 min, 1x/wk, 8 wks)^40^ or light stretching/strength training designed to match the same intensity as the Qigong group^41^ as parallel classes to the Qigong intervention. In contrast, our study required significantly more in-person instruction time for each group (2.25 hr classes, 2x/wk, 10 wks) and included an Exercise/Nutrition group which incorporated not only moderate intensity aerobic exercise, which has previously been demonstrated to improve fatigue ^16^, but also moderate-high intensity resistance training. This combination of exercise alongside a plant based diet and overall health counseling, all of which have also shown previous evidence in improving fatigue^15,18^, constituted an incredibly strong intervention group to compare Qigong against. The fact that the Qigong group improved fatigue levels by a similar amount as the Exercise/Nutrition group is therefore novel and provides an important baseline for future investigation. It further lends support to the finding of a recent meta-analysis of Qigong’s efficacy in fatigue reduction that Qigong may constitute an effective alternative to exercise for improving fatigue in cancer survivors^27,35^.

An important aspect of the current study was the large time demands required for participation, which had a few notable effects. First, it impacted the dropout rates across both classes, as the majority of participants who dropped cited too much time as their primary reason (See Figure 1, CONSORT flow chart). However, the retention rates in this study were still similar to those of other trails^40,63^. Second, those who completed had a fairly large improvement in fatigue regardless of group assignment, and this improvement could be in part due to the large amount of face-to-face instruction time (4.5 hrs, regardless of group, per week). This is much higher than many comparable studies, which typically include interventions meeting for 1.5-2 hours total in person (for example: McQuade at al 2017^41^, Larkey et al 2015^64^.

A critical component of behavioral interventions is the role of the teacher. Both intervention groups were taught by experts of their specific area: the Qigong group was taught by an internationally recognized master with more than 40 years experience and the Exercise/ Nutrition group was taught by a combination of behavioral change experts with licensing in nutrition and chiropractic medicine. The use of expert teachers, the large amount of in-person instruction, and social support provided from the group environment were therefore balanced across groups.

### 4.2 Development of Intervention Classes: National Institute of Health (NIH) Stage Model

While other mind-body interventions such as mindfulness have benefited from a replicable, manualized mindfulness program (eg Mindfulness Based Stress Reduction) that has followed an iterative process of testing and refinement, this has not been as robust among Qigong programs other than an existing generalized Qigong/Tai Chi Easy course^64,65^. Development and validation of effective behavior change interventions has been a target of the NIH stage model and a broader focus of the NIH Science of Behavioral Change (SOBC) initiative^66,67^. The NIH stage model was developed to maximize the feasibility, engagement, and efficacy of the interventions in this study^66^. In this model, behavioral interventions are developed and refined through an iterative process aimed at closing the gap between traditional implementation science and basic science to establish moderators of efficacy while also identifying underlying mechanisms. The goal of this iterative process is to ensure the intervention is continuously refined until it has reached the ability to be taught and practiced in the community with real-world effectiveness^66^. We therefore have utilized the NIH stage model to aid the development of a hospital and research-clinic friendly manualized Qigong program in an attempt to begin bridging this gap.

Because the Qigong program was a novel program designed by the instructor, and the Pre-Train (exercise) and CHIP (nutrition/health education) programs were combined together for the first time as the Exercise/Nutrition groups, we ran an initial pilot study with fatigued female cancer survivors in 2016 in accordance with the NIH stage model. Following the recommendation of the stage model, we used verbal feedback from participants through-out the interventions, as well as observations of study personnel and the intervention instructors, to refine and hone the delivery of the interventions to the cohort in the 2017 study reported on here. Notable changes included moving from 3 days a week to 2 days, but preserving the total number of delivery hours (4.5 total in both years) and standardizing the delivery of the Qigong program to follow the same sequence in each class. While we effectively moved from Stage 1 [“Intervention Generation/Refinement”] to Stage 2 [“Efficacy in Research Clinics”] in the NIH stage model between the 2016 and 2017 studies, the results of this study allow us to revisit Stage 1 and further refine the interventions and basic science measures to further the development of these interventions. The current data highlight the need for future testing and refinement of interventions with larger sample sizes and intent-to-treat analyses that limit biases introduced by the current per-protocol analysis. Future goals include the development of a teacher training program to widen the access to classes and feasibility of study and generalization of teaching from experts to community leaders as well as refinement of intervention demands to maximize participant retention. A significant contribution of this study is the groundwork laid to develop such a Qigong intervention in accordance with this stage model.

### 4.3 Other factors to consider when directly comparing Qigong and Exercise/Nutrition

In our small sample the Exercise/Nutrition group had lower mean values at baseline than the Qigong group for all measures shown to have significant pre to post treatment differences. However, due to the large variance across subjects, none of the measures were significantly different between the two groups at baseline. In our case, the Exercise/Nutrition group was more fatigued at baseline (had a lower mean value), which is notable in light of a recent finding that the magnitude of fatigue reduction was largest for those most fatigued at baseline^35^. This could indicate that if the Qigong group had been as fatigued at baseline as the Exercise/Nutrition, the Qigong intervention may have yielded more fatigue reduction. Additionally there was a high degree of variability seen in the improvement scores in both groups (see Fig 2, Table 3), which indicates that while some people greatly benefited from their assigned intervention, others in the same group did not experience the same change in fatigue. Given this high variability was evident across both groups, it may suggest that some people may have done better in the other group. It is also notable that the average age of the Qigong group (60.8 years +/- 5.7) was higher than the exercise/nutrition group (54.2 years +/- 10.3), and while this wasn’t significantly different it had a trend towards significance (p=.062). Future work is needed to understand if subpopulations of fatigued patients may be more responsive to Qigong vs exercise/nutrition. However, efforts towards personalization may be beyond what is feasible in a randomized control trial.

A considerable aspect of the current study is that Qigong, a practice that derives from Chinese medical/philosophical theory, was compared to standard exercise/nutrition practices predominant in Western culture. Women who were raised in the United States and had no active practice or knowledge of Qigong were the main participants. As such, the women randomized to the Qigong group may have had more to learn about the overall framework of Qi and how it related to concepts of health improvement than those randomized to Exercise/Nutrition, which are both commonly accepted to improve health. Women in the Exercise/Nutrition group may therefore have had a stronger expectation of fatigue improvement than those in the Qigong group. Accordingly, cultural background and expectation of fatigue relief may have played a role in early adoption of the classes, though it seems to have been surmounted given the similar improvements in fatigue across groups. This further suggests that Qigong’s practices are efficacious for fatigue relief even in people who were not raised in geographic regions where Qigong is culturally endemic and widely recognized. However, this may be driven largely by having a highly trained teacher who was able to contextualize and translate the practices across the specific cultural backgrounds of the participants. Future work is needed to more directly explore these components.

### 4.3 Possible Shared and Divergent Mechanisms of Fatigue reduction across Qigong vs Exercise/Nutrition training

Findings in our secondary exploratory post-hoc analysis revealed significant pre-post changes for the Qigong intervention in emotional distress, mood regulation, and perceived stress (see Table 3 for Significant Changes in Qigong group: POMS total mood disturbance, POMS confusion, POMS fatigue, POMS tension, DERS Clarity *trend*, DERS Goals, DERS Impulse, DERS Total and PHQ-9. For Trends toward significant changes see: DERS Clarity, DERS Nonacceptance,, DERS Total, PSS, PHQ-9), while the exercise/nutrition intervention was associated with significant improvements pre-post in fatigue and sleep metrics (see Table 3 for significant changes in Exercise/Nutrition Group: FSI Avg Severity, FSI # of days fatigued, PSQI subjective sleep quality, PSQI daytime dysfunction. For trends toward significance see: FSI composite, PSQI subjective sleep quality, PSQI daytime dysfunction, PSQI global). Such findings could be indicative of divergent mechanisms related to primary outcome of fatigue reduction (See Table 3 FACIT Additional Concerns) or overall quality of life improvement between the two interventions, which requires further study.

The physically fatiguing aspects of the Exercise/Nutrition class required from muscular exertion and increases in cardiorespiratory and metabolic demands during class may be partly responsible for the improvements in sleep/fatigue that showed significant change in this class^68^. It has been suggested that exercise interventions may promote fatigue recovery by improving muscular strength and metabolic and oxygen usage by the muscles^69^. The incorporation of a plant based diet in this Exercise/Nutrition class may further have facilitated fatigue reduction by improving nutritional status and nutrients available for muscular exertion and recovery^70^. While this combined intervention was effective for fatigue reduction in our sample, moderate-vigorous intensity exercise can be prohibitively intense

While the Qigong class did not focus explicitly on any components of psychological health or mental health education, it was effective in improving these metrics. The general practice typically emphasizes an intentional focus on linking breath and bodily movement to circulate Qi in the body, while maintaining a meditative awareness on sensations generated by the body during breath and movement (Liu and Chen, 2010 [Book]). As such, Qigong practice naturally incorporates movement with breath and meditation practice, potentially fostering a strengthening of physical form alongside a reduction of stress and clarity of the mind. This potentially indicates a more affective or stress-based mechanism of fatigue reduction from Qigong training. This interpretation is consistent with prior findings in Qigong and Tai Chi studies showing their efficacy in improving mood regulation, depressive symptoms, and stress^27,28,63,71–78^, and cognitive function^37^. Importantly, Qigong/Taiji movement alone without the specific mindset/attentional quality (eg sham Qigong) failed to improve fatigue as much as standard Qigong/Taiji at post intervention and 3 month follow-up^64^. This suggests the specific type of awareness cultivated in Qigong training in addition to the movement is important in fatigue reduction. Given that these were self-reported changes, future studies should pay attention to potentially divergent physiologic mechanisms by which Qigong vs Exercise/Nutrition interventions may alleviate fatigue and investigate potential objective measures associated with such mechanisms.

## 5. Study limitations and future trials

Though one strength of this study was its single-blind randomized study design, which is notably absent in many studies of Qigong^27,74^, several limitations could be improved upon in future trials. One limitation is that fatigue was only assessed before and after the intervention, and long term follow up could be important for potentially capturing divergence in responses as a function of time. Additionally, this study compared two parallel intervention groups without including a treatment-as-usual group or no-treatment control, which would have been useful for testing the magnitude of fatigue change between intervention vs no intervention but was not feasible due to recruitment limitations. Further, given that exercise, nutrition, and health/psychoeducation were combined in one group in this study, future studies could compare the separate components’ efficacy for fatigue reduction against Qigong or other mind-body meditative movement interventions.

Due to the preliminary nature of the study and the loss-to-follow up of participants who dropped out, we were able to carry out only a per protocol analysis of those who completed the intervention. While this allows us to estimate the effect of adherence to the intervention, it potentially introduces levels of bias that undercut the randomization efforts and overestimates the effect size, so future studies should aim to include both per protocol and intent to treat analyses^57^. A major limitation of this paper is the small sample size, which makes us underpowered to conduct non-inferiority or equivalence analyses and further limits our power to detect true effects. This could also be resolved in future larger scale trials.

## Data Availability

Materials, deidentified data and analysis code are available by emailing the corresponding author.

## Acknowledgements

We wish to thank Camilla Moore, Greg Salguiero, and Katie Lester for acting as the instructors in the exercise/nutrition intervention, and Harrison Moretz for developing and instructing the Qigong intervention. We wish to thank Brandan Cullen, Helen Ding, Nikisha Vaghjiani, Sophie Sandweiss, Sam Fredericks, Danielle Silva, Christopher Black, Juan Santoyo, Uday Agrawal, Arison Than and Brandon Cullen for their contributions to the data collection aspects of this study. We wish to thank Sara Lazar and Ellen Flynn for their professional input at different stages of the study.

