## Supplemental Table 1 for "The Vitality Project: A Randomized Control Trial Comparing Effects of Qigong and Exercise/Nutrition Training on Fatigue, Emotional Health, and Stress in Fatigued Female Cancer Survivors"

Supplementary Material

**Table S1**

**ANOVA results for questionnaire subscales with significant main effect of ‘Time’**

| **Questionnaire** | **DFn** | **DFd** | **SSn** | **SSd** | **F** | **P** | **Ges Effect Size:** |
| --- | --- | --- | --- | --- | --- | --- | --- |
| FACIT-F_AC | 1 | 22 | 754.522 | 1395.102 | 11.898 | .002 | .116 |
| FACIT-F_PWB | 1 | 22 | 104.805 | 270.888 | 8.512 | .008 | 0.0960 |
| FACIT-F_EWB | 1 | 22 | 37.893 | 153.357 | 5.436 | .029 | 0.061 |
| FACIT-F_Total | 1 | 22 | 3104.735 | 6890.618 | 9.913 | .005 | 0.101 |
| FACT-G | 1 | 22 | 798.152 | 2455.805 | 7.150 | .0140 | .074 |
| FACIT-F TOI | 1 | 22 | 1949.348 | 4675.595 | 9.172 | .006 | 0.100 |
| FSI_most severe | 1 | 22 | 19.837 | 67.476 | 6.468 | .0190 | .0620 |
| FSI_Average | 1 | 22 | 27.063 | 77.916 | 7.641 | .0110 | 0.114 |
| FSI_composite | 1 | 22 | 17.983 | 51.008 | 7.756 | .0.110 | 0.104 |
| FSI_ number of days fatigued | 1 | 22 | 23.544 | 33.706 | 15.367 | .0007 | 0.092 |
| FSI_% of day fatigued | 1 | 22 | 17.847 | 74.070 | 5.301 | .0310 | 0.059 |
| FSI_Global | 1 | 22 | 2000.938 | 7312.979 | 6.020 | .0230 | .077 |
| PSQI_Subjective Sleep Quality | 1 | 17 | 4.352 | 7.911 | 9.352 | .0070 | 0.131 |
| PSQI_Daytime Dysfunction | 1 | 17 | 1.601 | 4.978 | 5.468 | 0.032 | .041 |
| PSQI_Global | 1 | 17 | 65.695 | 151.2 | 7.386 | .0150 | .109 |
| MAIA_ Body Listening | 1 | 22 | 10.962 | 16.816 | 14.342 | .001 | .139 |
| MAIA_Emotional Awareness | 1 | 22 | 3.646 | 18.593 | 4.314 | .050 | 0.053 |
| MAIA_Self Regulation | 1 | 22 | 8.567 | 27.115 | 6.951 | .0150 | 0.111 |
| MAIA_Trusting | 1 | 22 | 3.761 | 16.454 | 5.029 | .0350 | 0.045 |
| POMS_ TMD | 1 | 22 | 3959.116 | 12883.36 | 6.761 | .0160 | 0.091 |
| POMS_ anger | 1 | 22 | 65.595 | 270.385 | 5.337 | .0310 | 0.058 |
| POMS_confusion | 1 | 22 | 64.040 | 242.273 | 5.815 | .0250 | 0.084 |
| POMS_fatigue/energy | 1 | 22 | 165.553 | 581.364 | 6.265 | .0200 | 0.087 |
| POMS_tension | 1 | 22 | 180.798 | 418.119 | 9.513 | .0050 | 0.106 |
| DERS_ Awareness | 1 | 22 | 50.714 | 211.203 | 5.283 | .0310 | 0.058 |
| DERS_ Goals | 1 | 22 | 22.273 | 70.706 | 6.930 | .0150 | 0.051 |
| DERS_ Impulse | 1 | 22 | 22.846 | 55.154 | 9.113 | .006 | 0.063 |
| DERS_strategies | 1 | 22 | 35.266 | 114.734 | 6.762 | .0160 | 0.0480 |
| DERS_total | 1 | 22 | 993.960 | 2190.706 | 9.982 | .005 | 0.098 |
| PSS | 1 | 22 | 70.573 | 207.427 | 7.485 | .0120 | 0.094 |
| PHQ9_depression | 1 | 22 | 94.409 | 212.070 | 9.794 | .005 | 0.114 |
| PHQ15_somatic symptom severity | 1 | 22 | 87.730 | 157.937 | 12.220 | .002 | 0.093 |
| Unmitigated Communion | 1 | 22 | 47.833 | 240.979 | 4.367 | .0480 | 0.029 |
| SF36_emotional limitations | 1 | 22 | 8194.930 | 25485.62 | 7.074 | .0140 | 0.1330 |
| SF36_emotional limitations (group effect) | 1 | 22 | 9875.842 | 27878.79 | 7.793 | .0110 | 0.1560 |
| SF36_fatigue | 1 | 22 | 4033.382 | 5267.949 | 16.844 | .000468 | 0.132 |
| SF36_ Emotional Well-Being | 1 | 22 | 1846.387 | 3976.280 | 10.216 | .004 | 0.144 |

All subscales that showed a significant difference in mixed effect ANOVA group x time interactions are reported here. All subscales reported here demonstrated a significant main effect for ‘time’ alone, unless denoted as having a significant main effect for ‘group’. Ges=generalized eta squared effect sizes.
